# Hardness of Herd Immunity and Success Probability of Quarantine Measures: A Branching Process Approach

**DOI:** 10.1101/2020.10.22.20216481

**Authors:** Sujit Kumar Nath

**Affiliations:** School of Computing, University of Leeds, LS2 9JT, Leeds, UK; Faculty of Biological Sciences, University of Leeds, LS2 9JT, Leeds, UK

**Keywords:** Age-dependent branching process, infectious disease, extinction probability, herd immunity, generation time distribution

## Abstract

Herd immunity refers to the collective resistance of a population against the spreading of an infection as an epidemic. Understanding the dependencies of herd immunity on various epidemiological parameters is of immense importance for strategizing control measures against an infection in a population. Using an age-dependent branching process model of infection propagation, we obtain interesting functional dependencies of herd immunity on the incubation period of the contagion, contact rate, and the probability of disease transmission from an infected to a susceptible individual. We show that herd immunity is difficult to achieve in case of a high incubation period of the contagion. We derive a method to quantify the success probabilities of quarantine measures to mitigate infection from a population, before achieving herd immunity. We provide a mechanistic derivation of the distribution of generation time from basic principles, which is of central importance to estimate the reproduction number *R*_0_, but has been assumed in an ad hoc manner in epidemiological studies, by far. This derivation of the generation time distribution has the generality to be applied in the study of many other age-dependent branching processes, such as the growth of bacterial colonies, various problems in evolutionary and population biology etc.

## I. INTRODUCTION

The propagation of an infectious disease in a population can be viewed as a branching process where an infected individual comes in contact with susceptible individuals in the population, and transmits the infection to the susceptible individuals with some probability of transmission. Branching process model of infection propagation has been widely used to study the propagation of HIV, Rabies etc. infections, both in the population, as well as in the cells or tissues of an infected individual [1–3]. There are many reasons for which a stochastic model of an infectious disease is much preferable than its deterministic counterparts [4, 5]. Due to the inherent randomness of the infectious diseases, arising from the incubation period of the contagions [6–9], infectious period of the infected individuals [10–13], contact and transmission probabilities etc. [14–16], a stochastic model is preferable than a deterministic one[17–20]. However, as all the models have their own advantages and limitations, both of the stochastic [18, 21] and deterministic [22] models, and sometimes a combination of the two [23], are used to model infectious diseases, as per their suitability.

Here we model the propagation of an infectious disease in a large homogeneous population as an age-dependent branching process. Like every other model, this simplified model also has limitations, and sometimes could be inappropriate to capture the detailed dynamics of some specific disease. However, our goal here is to present some new interesting theoretical understanding of the disease propagation, and the effects of interventions. We derive a few general theorems on age-dependent branching process, and apply them to study our model of infectious disease, to quantitatively understand the difficulty for a population to achieve herd immunity, depending on the incubation period of the contagion, social interactions, and transmission probability of the contagion from an infected to a susceptible individual. We also provide a quantitative method to estimate success probabilities of quarantine measures, depending on the epidemiological state of a population.

The distribution of generation time or serial interval time for an infectious disease [24–27] is a very important quantity to understand the transmission potential, and to calculate the basic and effective reproduction number which is a key parameter to estimate the epi-demiological state of an infection in a population [28, 29]. Until now, the distribution of generation time is estimated by fitting gamma, lognormal, Weibull, or Gaussian distributions to the transmission pairs data [26, 30–32]. However, there is no formal derivation available in favour of the particular choice of these distributions, and hence, they are chosen heuristically. In this paper, a general theorem on generation time is derived which can have large applicability in the study of infectious diseases, as well as any kind of age-dependent population dynamics, such as cell divisions [33, 34], growth of microbial colonies [35, 36] etc. We apply this general theorem to derive the distribution of generation time for our model of infectious disease.

In section II we introduce our age-dependent model of infection propagation. Section III presents the derivation of the functional dependence of herd immunity threshold on the incubation period, contact and transmission probabilities, and fraction of susceptible population. We study the success probabilities of various quarantine measures, depending on the state of the infection in the population, and compare early vs. later lockdown in a hypothetical population in sections IV and V, respectively. In section VI we prove a general theorem to derive the distribution of generation time from the generating function of the first generation progenies of any general age-dependent branching process, and apply it to derive the distribution of generation time for our model. We present a concise summary of this work in section VII.

## II. MODEL OF INFECTION PROPAGATION USING AN AGE-DEPENDENT BRANCHING PROCESS

To map the infection propagation with a branching process, we identify an infected individual to be the ancestor of the individuals to whom it transmits the infection directly. The individuals who are directly infected by an ancestor, are called the progenies of the first generation. The individuals who are infected directly by a progeny of the first generation, are called the progenies of the second generation, and so on. The set of all progenies or descendants of an individual is called a line. If the number of descendants at the *r*^*th*^ generation is zero for an individual, its line is called extinct at the *r*^*th*^ generation. Now, let an ancestor gives rise to *n* number of first generation progenies with probability *p*_*n*_, where *n* ∈ ℤ_+_, the set of all non-negative integers. Then the function

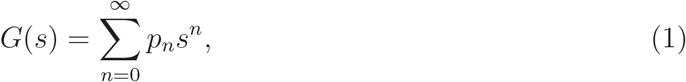

is the generating function for the distribution of number of progenies in the first generation. We now state a well-known theorem which establishes the basic reproduction number as a key parameter to determine the fate of an infection in a population, using the generating function of the first generation progenies.

### Theorem 1.

*Let G*(*s*) *be the generating function of a branching process for which the average number of progenies created by an ancestor be µ. If µ* ≤ 1, *the process dies out with probability one. If however, µ* > 1 *the probability x*_*r*_ *that the process terminates at or before the r*^*th*^ *generation tends to the unique root x* < 1 *of the equation s* = *G*(*s*), *as r* → ∞.

See [37, 38] for a proof of theorem 1.

In the infectious disease literature, the average number of first generation progenies (newly created infections) created by an ancestor (primary infected individual) is termed as basic reproduction number, symbolically denoted by *R*_0_ [28, 29]. Therefore, it is clear from theorem 1 that the infection will die out after sufficient time when *R*_0_ ≤ 1, which is called the herd immunity.

The assumptions of our model are as follows.

### Assumption 1

*The number of contacts made by an infected individual, with other individuals in the population, is a Poisson process with a rate α*.

### Assumption 2

*The population is large and homogeneous. Contacts made by the individuals in the population are independent of each other*.

### Assumption 3

*The rate of contacts is constant over time, unless any intervention is imposed, such as quarantine measures*.

### Assumption 4

*The incubation period and infectious period are used synonymously, because of the uncertainty of the initiation of the infectiousness in an individual carrying the contagion, and there is no relapse of infection*.

### Assumption 5

*Incubation period is memoryless*.

### Assumption 6

*The fraction of susceptibles in the population varies very slowly with respect to the incubation/infectious time period*.

We now prove a general theorem to derive the generating function for the number of first generation progenies, created by an ancestor, in an age-dependent branching process; and apply it to calculate the number of infectious contacts made by an infected individual for our model.

### Theorem 2.

*Let X(t) be a continuous time stochastic process with atomic distribution* ℙ(*X*(*t*) = *n*) = *p*_*n*_(*t*), *and generating function G(s,t), where n is any non-negative integer. Let X*_1_(*t*) *be the process X*(*t*) *with survival time T which is a continuous non-negative random variable with distribution function F* (*t*) = ℙ(*T* ≤ *t*). *Then the generating function G*_1_(*s, t*) *of the process X*_1_(*t*) *is given by*

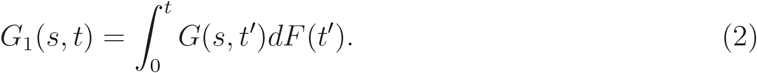

*Proof*. The given condition, ℙ(*X*(*t*) = *n*) = *p*_*n*_(*t*), implies the probability that *X*(*t*) = *n*, when the process survives at least for time *t*, is *p*_*n*_(*t*). Since the survival time *T* is distributed as *F* (*t*), the probability that *X*_1_(*t*) = *n* is

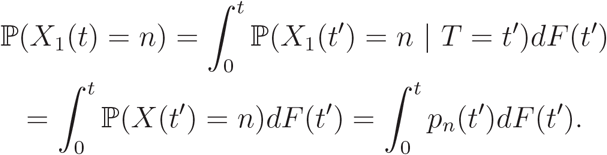

Therefore, the generating function for *X*_1_(*t*) is

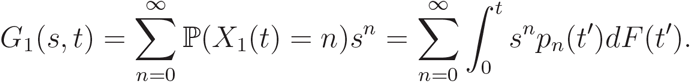

Now, identifying

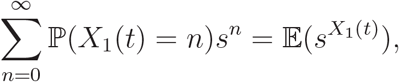

as an expectation value on a discrete/atomic measure, and applying Fubini’s theorem for changing the order of summation and integration, we obtain

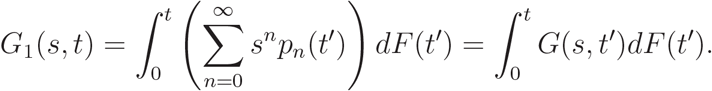

□

An important point to note here is that, in general, the function *G*_1_(*s, t*) could be the generating function of an improper distribution [39], i.e., *G*_1_(1, *t*) ≤ 1. In other words, the total probability that the number of progenies will be *n* = 0, 1, 2, 3, … etc., up to time *t*, could be less than or equal to 1.

### Corollary 3.

*If a Poisson process with rate α has exponentially distributed survival time with mean λ, then the generating function for the process at time t is given by*

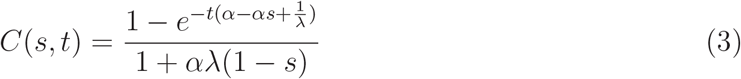

*Proof*. A Poisson process with rate *α*, has the generating function

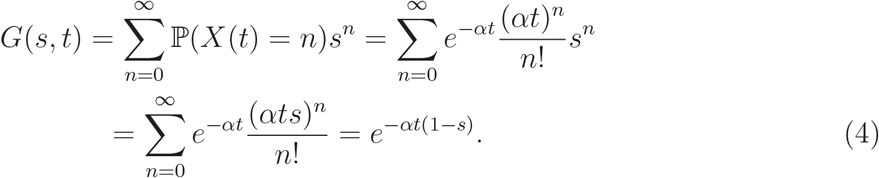

Using theorem 2 and equation 4, we therefore obtain the generating function for a Poisson process with exponentially distributed survival time having mean *λ* as

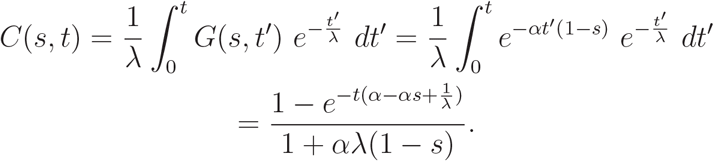

□

### Remark 4.

*Since* |*s*| *<* 1 *and α, λ* > 0, *we have* 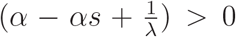. *Therefore, taking limit t* → ∞, *in equation (3), we obtain the generating function for the process, described in corollary 3, at the steady state as*

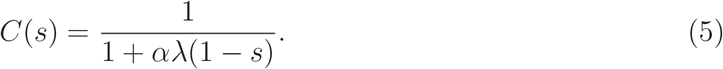

## III. HARDNESS OF HERD IMMUNITY

In our model of infection dynamics, an infected individual makes contacts with other individuals as a Poisson process with rate *α*, and remains infectious for an exponentially distributed random time period, having mean *λ*. Therefore, the generating function for the number of infectious contacts (contacts made while the person is still infectious) made by an infected individual up to time *t*, and during its full lifetime are given by *C*(*s, t*) and *C*(*s*) in equations (3) and (5) respectively.

We now recall a theorem for deriving the generating functions for compound distributions, which we shall use for some of our calculations.

### Theorem 5.

*Let N be a non-negative integral valued random variable with generating function G(s) and let* {*X*_*i*_} *be a sequence of independent and identically distributed (iid) non-negative integral valued random variables with generating function R*(*s*). *Then the generating function for the compound random variable S*_*N*_ = *X*_1_ + *X*_2_ + … + *X*_*N*_ *is G*(*R*(*s*)).

For a proof of theorem 5 see [37].

With our model assumptions, let us further assume that at time *t* the fraction of susceptibles present in a population be *p*_*s*_, and the probability of disease transmission from an infected individual to a susceptible individual, who have been in an infectious contact, be *p*_*c*_. Therefore, the probability of generating a new infected individual from a random contact (either with susceptible or with already infected/immuned) with an infected individual is *p*_*s*_*p*_*c*_. This event of creating a new infected individual, from a random contact with an infected individual, can be thought of as a Bernoulli trial *X* with success probability ℙ(*X* = 1) = *p*_*s*_*p*_*c*_ and ℙ(*X* = 0) = 1 − *p*_*s*_*p*_*c*_. The generating function corresponding to this Bernoulli trial is

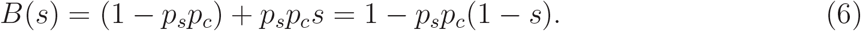

Now, the number of newly infected individuals from an existing infected individual, in time *t*, is the sum of *N* number of Bernoulli trials *X*, where *N* is a random number representing the number of infectious contacts made by the infected individual. Therefore, using theorem 5, from equation (3) and (6) we obtain the number of newly infected individuals, from an existing infected individual, in time *t* from the onset of its infection, has generating function

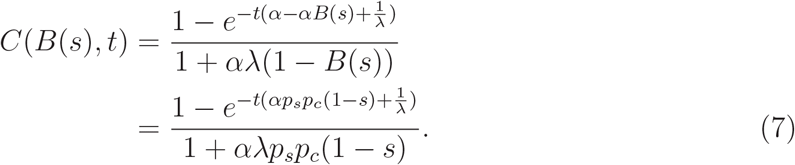

Taking limit *t* → ∞ in equation (7) we get the generating function for the same at steady state as

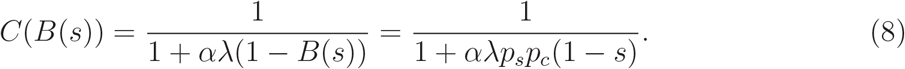

Equation (8) can also be obtained directly from equation (5), by replacing *s* by *B*(*s*) from equation (6). Note that here we are using assumption 6, i.e. the fraction of susceptibles *p*_*s*_, in the population, is constant over the infectious time period of an individual. Figure 1 shows the probability of the number of first generation progenies created by an infected individual, for different incubation periods of the contagion (evaluating the Taylor series coefficients of *C*(*B*(*s*)) in equation (8)). Equation (8) is the generating function for the number of newly created infected individuals, by an existing infected individual throughout its whole infectious period. Therefore, the average number of new infections generated by an existing infected individual is

**FIG. 1.**
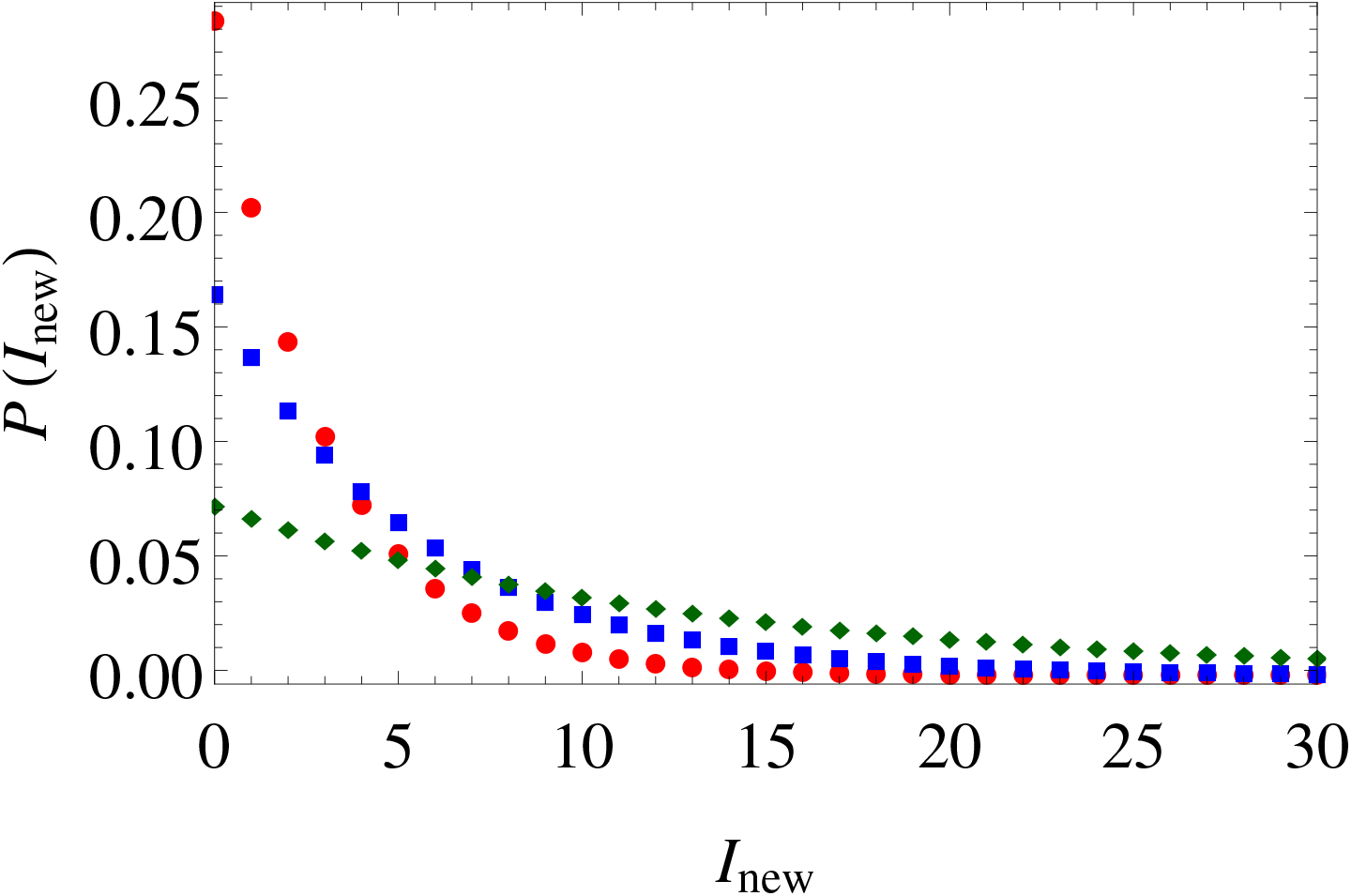
(Color online) Probability distributions of the number of newly created infections from an existing infected individual. Horizontal axis is represents the number of new infections created, and the vertical axis represents the corresponding probabilities. Red circle, blue square, and green diamond plots are for *λ* = 10, 20, and 50, respectively. *α* = 10, *p*_*s*_ = 0.5, *p*_*c*_ = 0.05 for all plots.

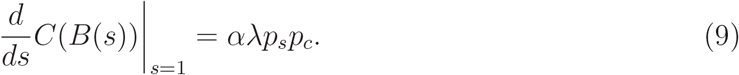

Recalling the definition of basic reproduction number *R*_0_ for an infectious disease, as the average number of new infections generated by an existing infected individual, we identify that equation (9) gives the *R*_0_ for our model of infection propagation. It is clear from equation (9) that *R*_0_ is a function of the fraction of susceptibles present in the population (*p*_*s*_), probability of disease transmission (*p*_*c*_), the rate of contacts between the individuals in the population (*α*), and the average incubation period (*λ*). Now, using theorem 1 we can conclude that the disease will die out over time, with probability one, if *R*_0_ = *αλp*_*s*_*p*_*c*_ ≤ 1, i.e. if

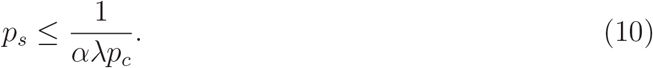

Inequality (10) has many significant implications about the achievement of herd immunity. First of all, it gives an upper bound for the fraction of susceptibles in the population, below which the infection in the population dies out with probability one, which is known as the herd immunity in epidemiological terminology. It is also evident from inequality (10) that when the mean incubation period (*λ*) is high, the upper bound of *p*_*s*_ is small. This means, to achieve herd immunity in the case of a contagion with high incubation period, (1 − *p*_*s*_) (which is close to 1) fraction of total population must be either immuned or infected, and hence, very hard to achieve. Proper use of personal protective equipments (PPE), like masks, hand sanitizers, face shields etc. can reduce the probability of disease transmission (*p*_*c*_), and hence helps to achieve herd immunity easily. Alternatively, imposing quarantine measures can reduce the rate of contacts between infected and susceptible population (*α*), and helps to achieve herd immunity. Therefore, in absense of any pharmaceutical interventions, like vaccines or other medicines, the alternative way to mitigate infection is by the use of PPE and/or quarantine measures.

## IV. SUCCESS PROBABILITY OF QUARANTINE MEASURES

Denoting the generating function for the number of first generation progenies created by an ancestor as *G*(*s*), we now recall a theorem for calculating the extinction probability of the line of a single ancestor.

### Theorem 6.

*For a branching process where the number of progenies produced by each particle are iid random variables, with generating function G*(*s*) *for the first generation progenies, the extinction probability of a line is given by the minimum of the positive roots of the equation x* = *G*(*x*), *and* 1.

See [37, 38] for a proof of theorem 6.

In our model of infection propagation, the generating function for the number of first generation progenies (newly generated infections) from a single infected individual is given by *C*(*B*(*s*)) in equation (8). Therefore, the extinction probability of all progenies (of all generations, i.e. the line of an ancestor) of an infected individual is obtained by solving the equation *x* = *C*(*B*(*x*)), which is same as the quadratic equation

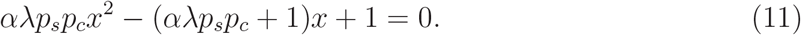

Here we make a strong use of assumption 6, which assumes that *p*_*s*_ varies so slowly that it remains almost constant for a line of an ancestor. This assumption can be supported by realizing that the line of an ancestor is likely to be in the same region or environment in the population. Equation (11) has two roots, 1 and 1*/αλp*_*s*_*p*_*c*_. Therefore, with the help of theorem 6, the extinction probability for all progenies or line of an infected individual is given by

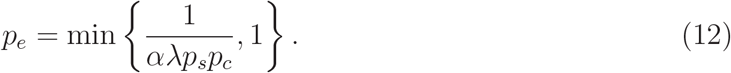

We now intend to calculate the number of individuals that will remain infected after time *t* = *T* if we start observing a fixed *N*_*I*_ number of infected individuals from time *t* = 0.

### Theorem 7.

*Let the incubation period T*_*I*_ *of a contagion has a distribution function* ℙ(*T*_*I*_ ≤ *t*) = *F* (*t*). *If we start observing N*_*I*_ *number of infected individuals, each infected from time t* = 0, *then the generating function for the number of individuals who will still remain infected after time t* = *T is given by*

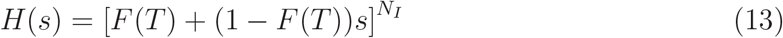

*Proof*. An infected individual remains infectious after time *T*, only when *T*_*I*_ *> T*. Since the incubation time *T*_*I*_ has distribution function *F* (*t*),

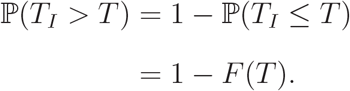

Now, we can think of *N*_*I*_ number of Bernoulli trials with success probability (1 − *F* (*T*)). The total number of successes of these Bernoulli trials results the number of individuals who will still remain infectious after time *T*. In other words, this number is given by the random variable 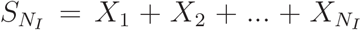, where each *X*_*i*_ is a Bernoulli random variable with ℙ(*X*_*i*_ = 1) = 1 − *F* (*T*), and ℙ(*X*_*i*_ = 0) = *F* (*T*). The generating function for each Bernoulli trial *X*_*i*_ is [*F* (*T*) + (1 − *F* (*T*))*s*]. Therefore, using theorem 5 we obtain the generating function of 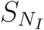 as

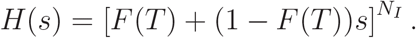

□

### Corollary 8.

*Let the incubation period of a contagion be exponentially distributed with mean λ. If we start observing N*_*I*_ *number of infected individuals from time t* = 0, *then the generating function for the number of individuals who will still remain infected after time t* = *T is given by*

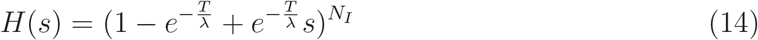

*Proof*. As the exponential distribution is memoryless, we need not worry about the instants when each individual got infected. Therefore, we can assume that all of the *N*_*I*_ individuals got the infection, simultaneously at *t* = 0, i.e. the time we start our observation. Since the incubation period *T*_*I*_ is exponentially distributed with mean *λ*, its distribution function is given by

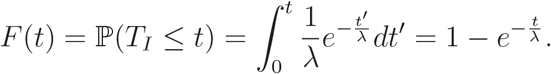

Therefore, using theorem 7, we obtain the desired result. □

Figure 2 shows the probabilities of the number of remaining infected individuals (among those who are being observed) after the time duration of observation, by plotting the Taylor series coefficients of equation (14).

**FIG. 2.**
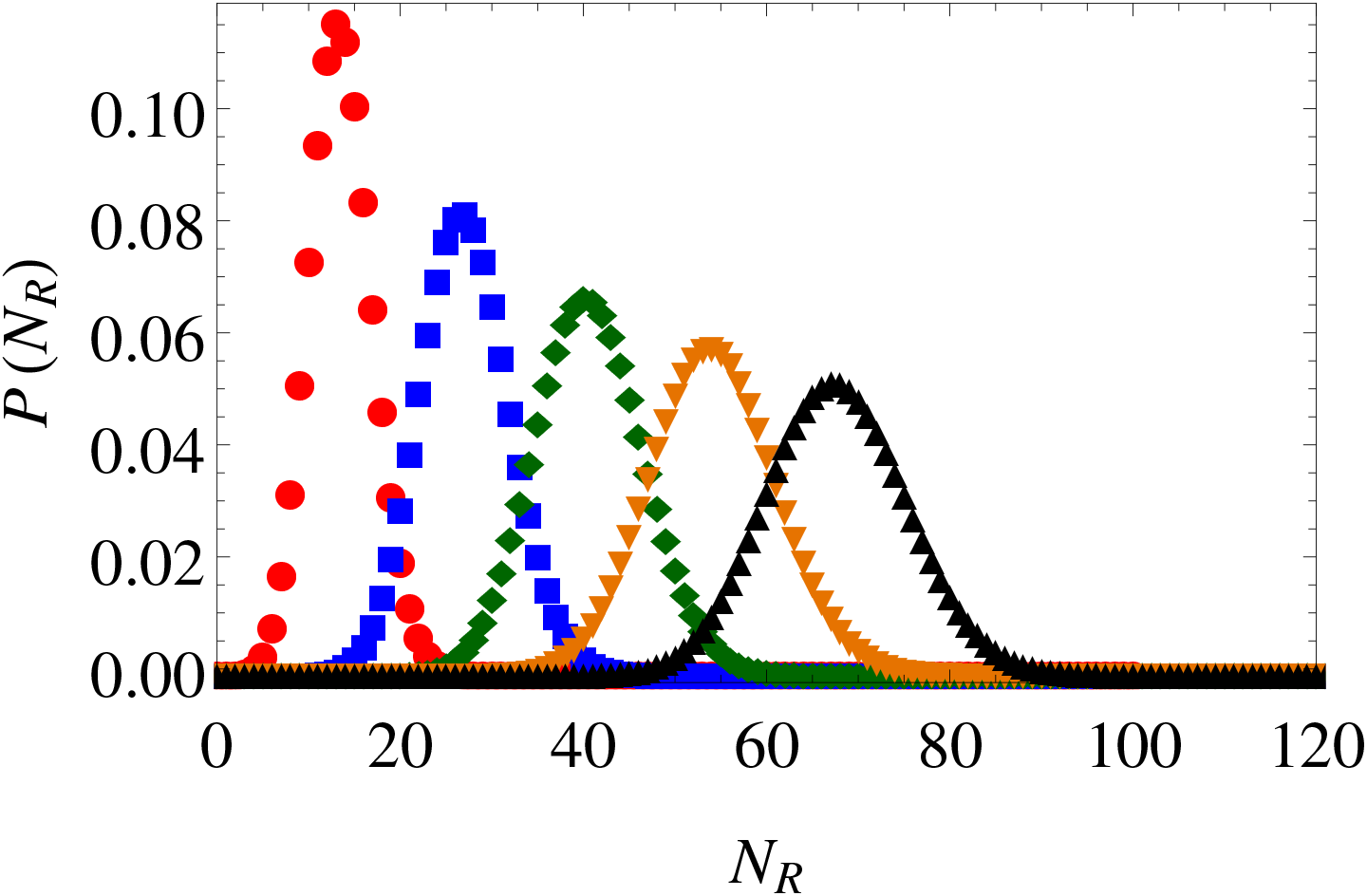
(Color online) Probability distribution for the number of infected individuals remaining after the withdrawal of quarantine measure. The horizontal axis denotes the number of infected, remaining after *T* = 40 days of quarantine. The average incubation period *λ*, of the contagion, is taken to be 20 days. The number of infected *N*_*I*_ at time *t* = 0 is taken to be 100, 200, 300, 400, and 500, for the red circle, blue square, green diamond, orange inverted triangle, and black triangle plots, respectively.

We now prove a theorem which will be used for calculating the extinction probability of the infectious disease by quarantine measures.

### Theorem 9.

*Let N number of iid Bernoulli trials, each having success probability p, are performed, where N is a positive integral valued random variable with generating function Q*(*s*). *Then the probability of obtaining all success is Q*(*p*).

*Proof*. Let ℙ(*N* = *k*) = *q*_*k*_, for *k* = 0, 1, 2, Then by the definition of generating function

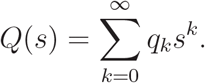

By conditioning on the number of trials *N*, we obtain the probability of all success in random number of Bernoulli trials as

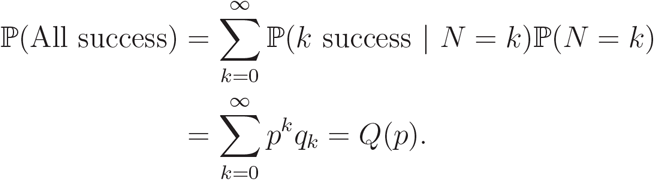

□

Theorems 7 and 9 together can be used to estimate the success probability of any quarantine measure in mitigating infection. Here we assume an ideal quarantine measure where no contact between an infected person and a susceptible person occurs. Let at some point of time there be *N*_*I*_ number of infected individuals present in the population. Let an ideal quarantine measure is imposed at that time for next *T* days, and after *T* days the quarantine is withdrawn. During the quarantine period *T*, some infected individuals can be recovered and others will remain infected. The generating function for the number of individuals (*N*_*R*_), who will still remain infectious after *T* days of quarantine, is given by *H*(*s*) in equation (14). After the quarantine is withdrawn, the remaining infected individuals will again start interacting with the susceptible individuals in the population and create new infections. The probability that the number of all progenies of a single infected individual will be zero, after sufficient time, is given by *p*_*e*_ in equation (12). Now, the event that the total number of progenies of all the remaining *N*_*R*_ number of infected individuals is zero after sufficient time, is equivalent to the event that *N*_*R*_ number of iid Bernoulli trials, each having success probability *p*_*e*_, results all successes. Since *N*_*R*_ has the generating function same as in equation (14), using theorem 9, we obtain the probability that the infection will be mitigated after sufficient time from the withdrawal of quarantine is

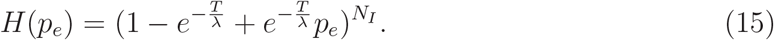

As *T* → ∞ in equation (15), the extinction probability *H*(*p*_*e*_) → 1. This implies that as the duration of quarantine increases, the probability of infection mitigation becomes higher. Also, when *p*_*e*_ = 1, i.e. *αλp*_*s*_*p*_*c*_ ≤ 1 (by equation (12)), and hence herd immunity is achieved, *H*(*p*_*e*_) = 1, i.e. the infection will be mitigated with probability one. Figure 3 shows the probability of zero infection as a function of the duration of quarantine, before achieving herd immunity.

**FIG. 3.**
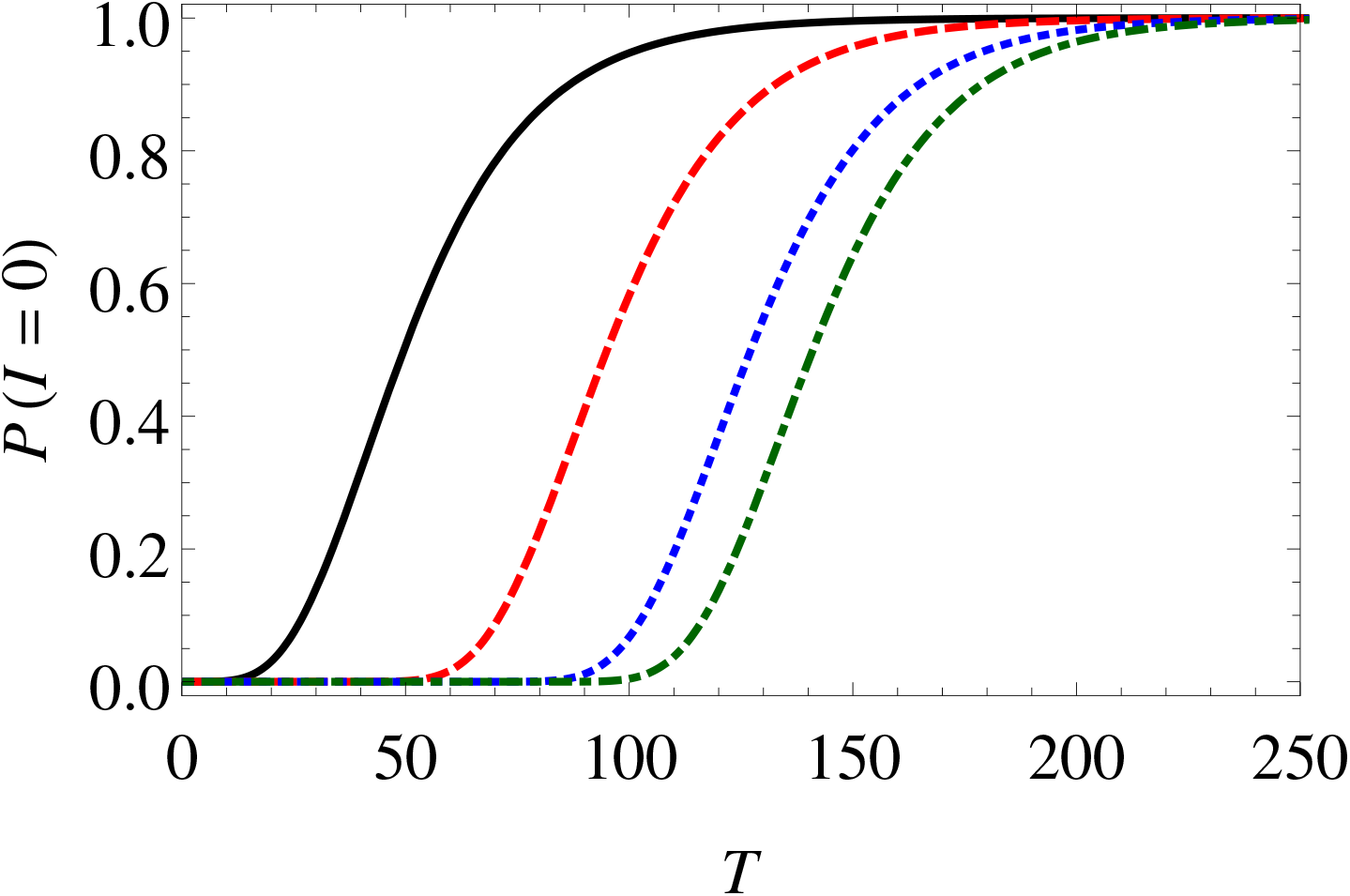
(Color online) Extinction probabilities of the infection as a function of the duration of quarantine. Horizontal axis represents the duration of quarantine, and vertical axis represents the corresponding extinction probabilities. Black solid, red dashed, blue dotted, and green dot-dashed curves are for *N*_*I*_ = 10, 100, 500, and 1000 respectively. *α* = 10, *λ* = 20, *p*_*s*_ = 0.5, *p*_*c*_ = 0.05 are taken for all plots.

## V. EARLY IMPOSITION OF LOCKDOWN CAN SOMETIMES BE LESS EFFECTIVE THAN A DELAYED IMPOSITION

We now study our infection propagation model in a toy population, and see some counter-intuitive results. Let the size of the total population be *N* = 10000, among which *N*_*I*_ number of individuals are infected at time *t* = 0. Therefore, the number of susceptibles at time *t* = 0 is *N* − *N*_*I*_. Hence, the fraction of susceptibles in the population is *p*_*s*_ = (*N* − *N*_*I*_)*/N*. Let the transmission probability *p*_*c*_ be 0.05. Let us consider that there is no relapse of infection, i.e., if someone has ever been infected, they cannot be susceptible anymore. Consequently, the fraction of susceptibles cannot be higher than (*N* −*N*_*I*_)*/N* for any *t* ≥ 0. If we now calculate the extinction probability as in equation (15), as a function of lockdown/quarantine duration, we obtain Figure 4. We see in Figure 4 that when *N*_*I*_ = 8500, the infection dies out with higher probability than the situation when *N*_*I*_ = 5000 or 6000, upon the imposition of quarantine measures of same duration. It is to be noted that none of these three cases have achieved herd immunity threshold, as *αλp*_*s*_*p*_*c*_ *>* 1 for all three cases. The reason for this is, as the size of the infected (or immune, after recovery from infection) population increases, the fraction of susceptibles *p*_*s*_ decreases, and so is the probability of infectious contacts with susceptibles. Hence, it is a tug of war between the number of infected *N*_*I*_, and the fraction of remaining susceptibles *p*_*s*_ = (*N* − *N*_*I*_)*/N*, in the population, to maximize the probability of total extinction, *H*(*p*_*e*_) in equation (15).

**FIG. 4.**
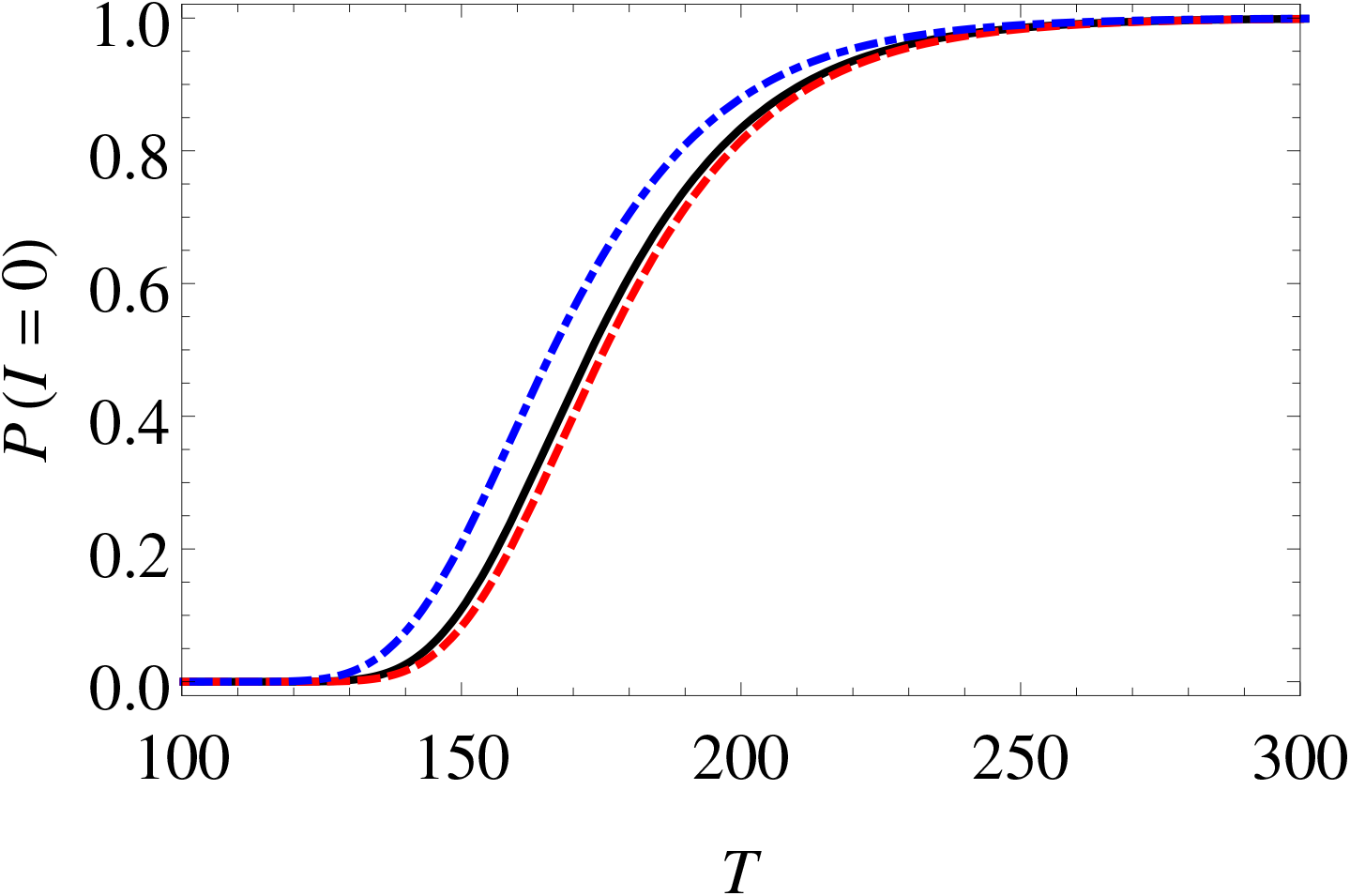
(Color online) Extinction probabilities of the infection as a function of the duration of quarantine, in a toy population of size *N* = 10000. Horizontal axis represents the duration of quarantine, and vertical axis represents the corresponding extinction probabilities. Black solid, red dashed, and blue dot-dashed curves are for *N*_*I*_ = 5000, 6000, and 8500, respectively. *α* = 10, *λ* = 20, *p*_*c*_ = 0.05 are taken for all plots.

This, therefore, implies that imposing an early quarantine measure may not always be most effective, unless the quarantine is maintained sufficiently long. Otherwise, it is a better strategy to let the infection spread into the population up to some time to ensure the number of remaining susceptibles is relatively low, and then impose the quarantine measure for the same duration. The later strategy will then have a higher probability of success in mitigating infection from the population.

It is to be noted here that after the withdrawal of lockdown the infection still keeps on propagating in the population to some extent, even before eradication. Therefore, it needs a detailed calculation to comment on the total number of infections occurring before it is eradicated from the population (known as the final size of infection, in the literature) [17, 18]. However, it is also to be understood that when total *N*_*I*_ number of people become infected, up to some point of time from the initiation of the infection in the population, not all of them remains infected at that time instant since some of them have already recovered and have become immune to the infection. So, the extinction probabilities in figure 4 is prone to underestimation. Therefore, to decide on the best strategy for mitigating an infection from a population, using quarantine measures, it needs an extensive analysis, to save both economy and healthcare system in an optimized way [40].

## VI. THE DISTRIBUTION OF GENERATION TIME

The generation time for an infectious disease is defined as the time interval between the onset of infection in an individual to the first generation of another new infected individual by the primary infected individual [24]. We now calculate the distribution of the generation time for our model of infectious disease. To derive the distribution function of the generation time we need to use the generating function for the tail of a discrete random variable. More specifically, let *N* be a non-negative integral valued random variable having distribution ℙ(*N* = *k*) = *p*_*k*_ (possibly improper distribution), for *k* ∈ {0, 1, 2, …}. Let *M* be defined as the tail of *N* having distribution ℙ(*M* = *k*) = ℙ(*N > k*) = *p*_*k*+1_ + *p*_*k*+2_ + … = *q*_*k*_, for *k* ∈ {0, 1, 2, …}. We now state a theorem which relates the generating functions of *N* and *M*.

### Theorem 10.

*If the generating function of N is P* (*s*), *then the generating function for its tail M is given by*

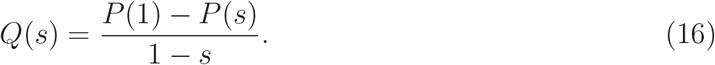

*Proof*. The proof goes exactly in the same line as in [37], where the generating function for the tail distribution is derived for a proper distribution. By definition,

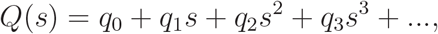

where *q*_*k*_ = *p*_*k*+1_ +*p*_*k*+2_ +…, for *k* ∈ {0, 1, 2, …}. Therefore, the coefficient of *s*^*n*^ in (1−*s*)*Q*(*s*) equals *q*_*n*_ − *q*_*n*−1_ = −*p*_*n*_ when *n* ≥ 1, and equals *q*_0_ = *p*_1_ + *p*_2_ + *p*_3_ + … = *P* (1) − *p*_0_ when *n* = 0. Therefore,

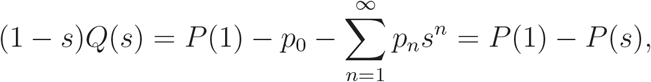

and hence the desired result. □

We now derive a general theorem to calculate the distribution of generation time, with the help of theorem 10.

### Theorem 11

(Generation time distribution). *Let G*(*s, t*) *be the generating function for the number of first generation progenies, created by an ancestor of an age-dependent branching process up to time t, with steady state generating function G*(*s*), *i*.*e*. lim_*t*→∞_ *G*(*s, t*) = *G*(*s*). *Let the number of progenies created by the ancestor throughout its lifetime be greater than or equal to m. Then the time τ*_*m*_, *required to create the first m number of progenies by the ancestor, has the distribution*

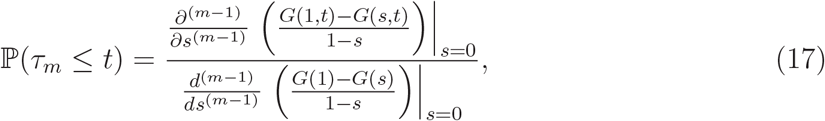

*and hence, the generation time τ has the distribution*

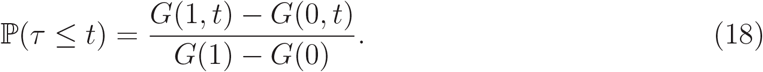

*Proof*. The event that *τ*_*m*_ ≤ *t* is same as the event that the number of progenies *N* (*t*), created up to time *t*, is greater than or equal to *m*. This implies

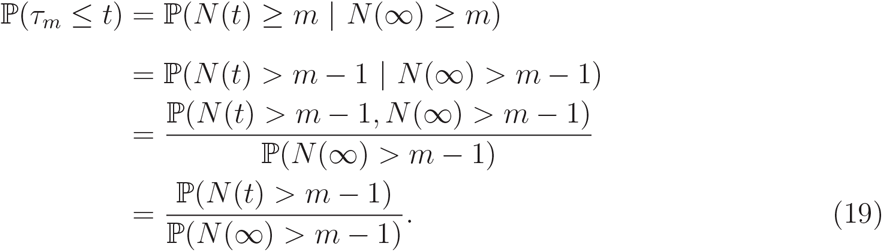

Identifying ℙ(*N* (*t*) *> m* − 1) as the tail distribution of the branching process under consideration, at time *t*, using theorem 10 we conclude that its generating function has the form

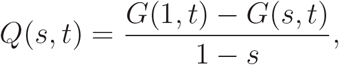

and ℙ(*N* (*t*) *> m* − 1) is the coefficient of *s*^*m*−1^ of the Taylor series expansion of *Q*(*s, t*) with respect to *s*. Similarly, ℙ(*N* (∞) *> m* − 1) is the coefficient of *s*^*m*−1^ in the Taylor series expansion of *Q*(*s*), where *Q*(*s*) = lim_*t*→∞_ *Q*(*s, t*). Hence, substituting these values of ℙ(*N* (*t*) *> m* − 1) and ℙ(*N* (∞) *> m* − 1) in equation (19) we obtain equation (17). Finally, we obtain equation (18) from equation (17), for *m* = 1. □

Having obtained the general procedure to derive the distribution function of generation time, we now apply theorem 11 to derive the distribution of generation time for our model.

In our model we have *G*(*s, t*) = *C*(*B*(*s*), *t*). Therefore, using equations (7) and (18), we obtain the distribution function for generation time as

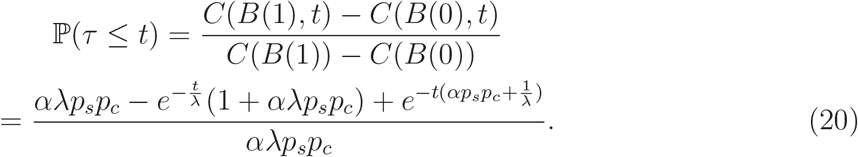

Therefore, the density function for the generation time is given by

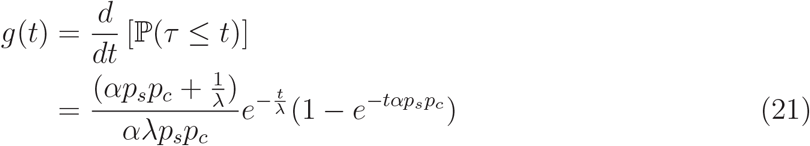

Figure 5 plots the probability density function *g*(*t*), for the generation time, in case of two different incubation periods. As seen from the plot that the mode of the distribution does not change much with the incubation period. Dependence on other parameters can also be checked, and it can be shown that decreasing *α, p*_*s*_, and *p*_*c*_ will shift the mode towards right, as it is evident that decreasing these parameters will delay the generation of a new infection.

**FIG. 5.**
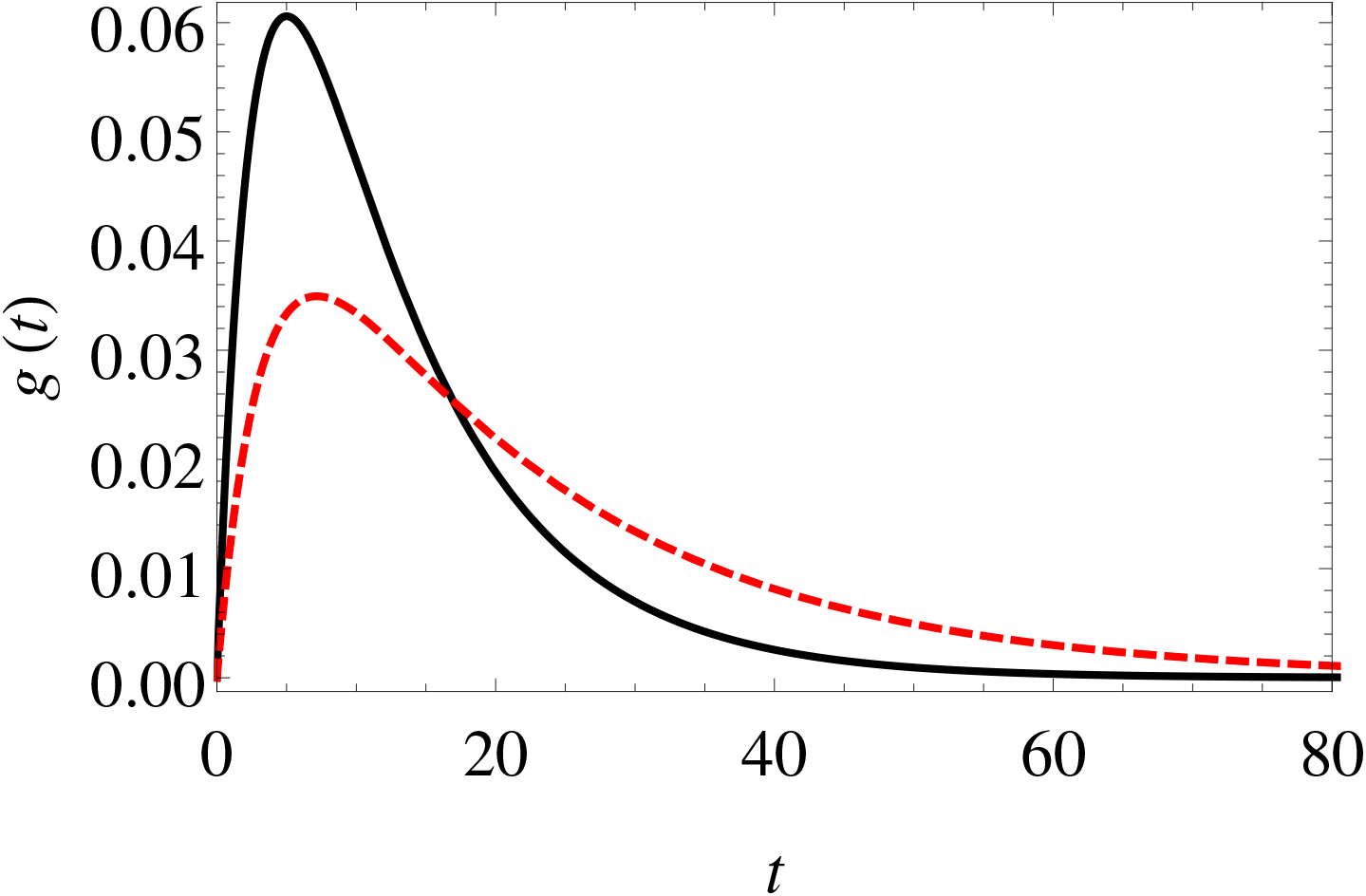
(Color online) Probability density function for the generation time. Black solid curve is for *λ* = 10; and red dashed curve is for *λ* = 20. Other parameters for both the curves are taken to be *α* = 10, *p*_*s*_ = 0.5, *p*_*c*_ = 0.05.

## VII. CONCLUSION

Modeling the propagation of an infectious disease, in a population, as an age-dependent branching process, we obtain the functional dependence of herd immunity on various epidemiological parameters. We show that herd immunity is difficult to achieve when the incubation period is high. We show that the mass use of PPE can help to achieve herd immunity, and to eradicate the infection faster from the population. We provide a method to estimate the success probabilities of various quarantine measures, and show by considering a hypothetical situation that an early imposition of lockdown may not always be a better strategy against a delayed imposition of lockdown of the same duration. We derive a general theorem to calculate the distribution of generation time, which can be used in the study of many systems, modeled as age-dependent branching process. Using this theorem we calculate the generation time distribution for our model of infection propagation, and obtain a two parameter (effectively) distribution which can be used in epidemiological studies, as a logical replacement of hitherto used heuristic distributions, such as gamma, lognormal, Weibull, Gaussian etc. [26, 27, 31, 32, 41].

## Data Availability

The manuscript uses no data files.

